# Analyses of Omicron genomes from India reveal BA.2 as a more transmissible variant

**DOI:** 10.1101/2022.04.25.22274272

**Authors:** Ashwin Atkulwar, Akif Rehman, Y. Imaan, Mumtaz Baig

## Abstract

This is the first study on omicron genomes from India to focus on phylodynamics and phylogenomics trait to provide an insight into the evolution of omicron variants. We analyzed 564 genomes deposited to GISAID database from various states of India. Pangolin COVID-19 Lineage Assigner tool was used to determine lineage assignment of all retrieved genomes. A Maximum likelihood (MLE) tree construction further confirms the separation of genomes into two distinct clades, BA. 1. and BA. 2. A very high reproduction number (R0) of 2.445 was estimated for the lineage BA.2. The highest R0 value in Telangana confirms the prevalence of lineage BA.2 in the state. Construction of the Reduced Median (RM) network shows evolution of some autochthonous haplogroups and haplotypes, which further supports the rapid evolution of omicron as compared to its previous variants. Phylogenomic analyses using maximum likelihood (ML) and RM show the potential for the emergence of sub-sublineages and novel haplogroups respectively. Due to the recombinant property and high transmissibility of omicron virus, we suggest continuous and more widespread genome sequencing in all states of India to track evolution of SARS-CoV-2 in real time.

## Introduction

Virus genomes display high diversity and are capable of adapting to a variety of hosts with a wide range of tissue tropism and thus increased likelihood of emergence of novel variants [1]. Through mutation, viruses continuously evolve into new variants. According to their severity, these variants may be classified under variants being monitored, variants of interest, variants of concern, and variants of high consequence. Following the report of the first case on December 31, 2019, WHO officially declared an outbreak of n-CoV-2 as a global pandemic on March 11, 2020. Five sub-variants (Alpha, Beta, Gamma, Delta, and Omicron) have been identified to date. A case of novel severe acute respiratory syndrome coronavirus 2 (SARS-CoV-2) B.1.1.529 is named as Omicron (O) has been reported from Botswana and South Africa on 11^th^ and 14^th^ November 2021, respectively [2] The variant of concern, omicron (B.1.1.529), is circulating across the globe with different sub-lineages including BA.1 (B.1.1.529.1), BA.2 (B.1.1.529.2) and BA.3 (B.1.1.529.3). In a report from WHO, omicron cases were confirmed in 149 countries, but current data suggest that this variant is detected in approximately 177 countries, so international travel is the most likely way for this variant to spread globally. The omicron gains attention due to the number of mutations in the spike protein gene, which influences its viral transmissibility, replication, and binding of antibodies, and its dramatic increase in South Africa [3]. Based on baseline studies, it was confirmed that the new variant was able to significantly escape immunity developed from previous infections and vaccinations [4, 5]. According to the WHO report and evidence from the studies, the risk associated with the omicron remains very high. Omicron shows major advantages over the Delta variant in terms of rapid community transmission, causing dramatic increase in the amount of cases in communities. Despite the lower risk of severe symptoms and deaths, omicron infection rate in many countries, including India, are very high, resulting in higher hospitalization rates and increased pressure on health care facilities. About 91,781 genomes were submitted to the GIASID database from around the world, out of which India shared 4283. Genome sequencing and annotation of novel SARS-CoV-2 variants from the different countries provide baseline data for investigating the molecular epidemiology of this continuously evolving virus [6]. With molecular epidemiology, it is possible to monitor the ongoing pandemic in relation to vaccine development. To determine the underlying phylogeographic and phylodynamic pattern of this omicron variant in the Indian context, we analyzed, 564 genomes deposited from different states of India. This study examined (i) different sub-variants circulating in different states (ii) their evolution within and outside states (iii) estimation of transmission with respect to reproduction number (R0) (iv) efforts of various states in genome sequencing in tracking the variants.

### Methodology

From the GISAID database, whole genome sequences for omicron (B.1.1.529) variants deposited until 29^th^ January 2022 from different states of India were retrieved. We analyzed only complete genomes with high coverage, while gaps and incomplete genomes with missing information were discarded. In total, 564 genomes were aligned using MAFT version 7 [7], and haplotype numbers were estimated using DnaSP v624 [8]. Prior to alignment, all 564 genomes were assigned lineages using Pangolin COVID-19 Lineage Assigner (https://pangolin.cog-uk.io/) [9]. A final dataset consisting of 297 haplotypes was compiled after excluding 7 genomes with many missing bases. A multi-type birth-death model was used in BEAST v2.6.6 [10], in which a HKY nucleotide substitution model with a gamma category count of 4, with a strict clock rate of 2.4E-4 was applied. Parameters were sampled in MCMC analysis every 1000 generations over a total of 10 million generations. BecomeUinfectiousRate and basic reproduction number (R0) were set with the mean distribution of 52.0, assuming a mean recovery time of 7 days. A number of parameters such as, effective sample size, 95% highest posterior density intervals, reproduction number (R0), BecomeUinfectiousRate were examined in Tracer v1.7.0 [11]. Based on 290 genomes from all major states of India, the Reduced Median Network (RM) was constructed using the program Network 10 [12]. Online version of PhyML3.0 was used to construct maximum likelihood tree using the same number of genomes [13].

## Results

### Haplotype estimation, lineage assignment and reproduction number (R0) estimation

After aligning 564 genomes, 297 haplotypes were estimated and used further for downstream analyses. As result of lineage assignment using Pangolin COVID-19 Lineage Assigner, 198 genomes were found to be BA.1, 92 genomes to be BA.2 and no BA.3 lineage was detected. For the BA.2 lineage, a reproduction number (R0) of 2.445 was found (HPD=1.525-3.761) (Figure 1). Reproduction number (R0) for lineage BA.1 was found to be 1.700 (HPD=1.143-2.460). Similarly, R0 values of omicron variants from various states in India range from 0.959 to 1.624 (Table 1 and Figure 1). The highest transmissibility was found in Telangana, which had the highest prevalence of BA.2 lineage in India, while the lowest reproduction rate (R0) was observed in Uttar Pradesh (UP) (Table 1).

**Figure 1.**
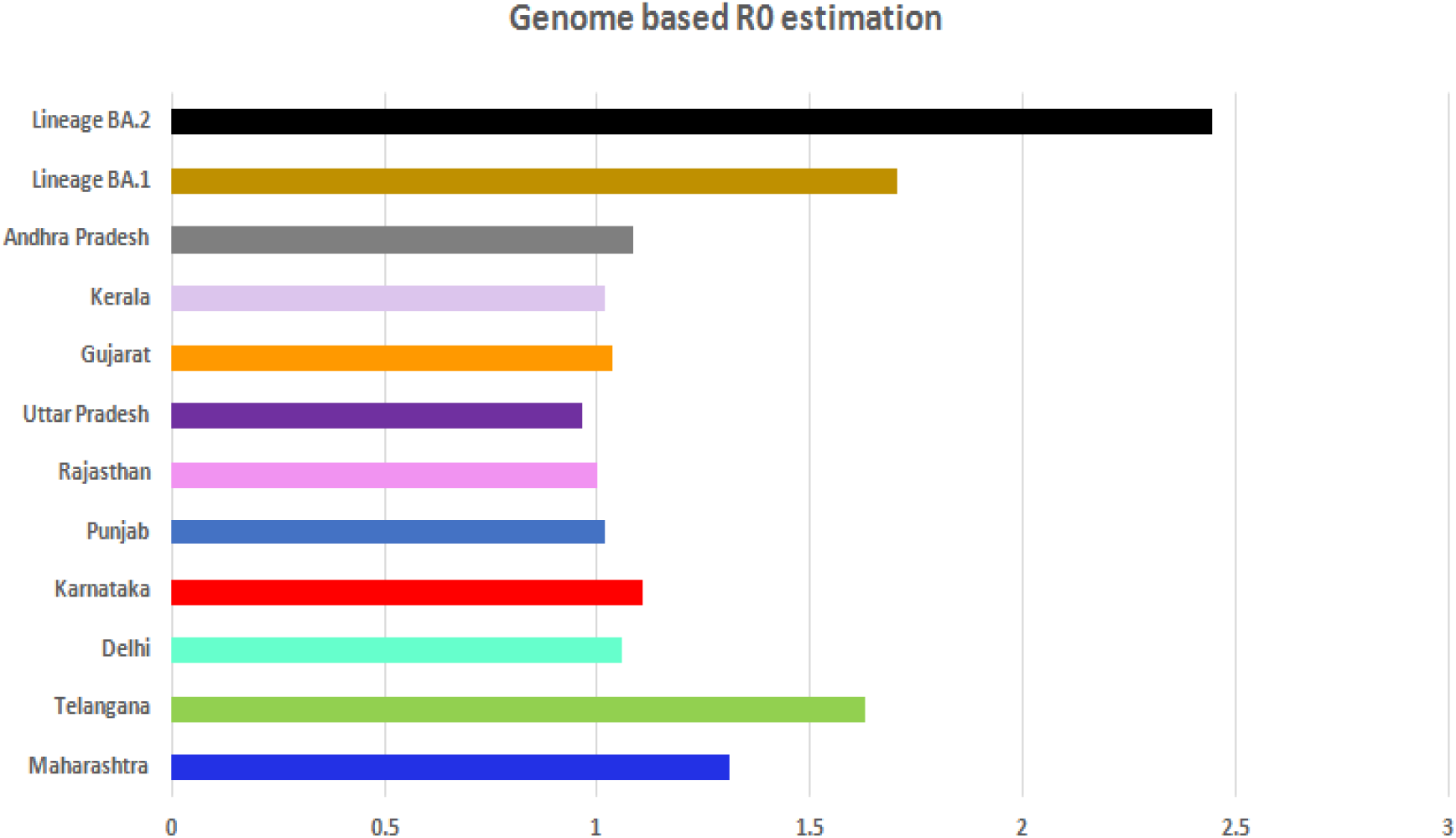
The basic reproduction number (R0), calculated for the various states of India. The graph also demonstrates the comparative analysis of R0’s of BA.1 and BA.2 omicron lineages.

**Table 1.** Comparative R0s of the omicron variants from different states of India with 95% HPD intervals.

| Sr. No. | State | R0 | 95 % HPD Interval |
| --- | --- | --- | --- |
| 1 | Uttar Pradesh | 0.959 | 0.171 – 1.526 |
| 2 | Kerala | 1.013 | 0.146 – 1.525 |
| 3 | Andhra Pradesh | 1.08 | 1.010 – 1.184 |
| 4 | Telangana | 1.765 | 1.226 – 2.695 |
| 5 | Delhi | 1.052 | 1.012 – 1.110 |
| 6 | Maharashtra | 1.306 | 0.996 – 1.734 |
| 7 | Gujarat | 1.033 | 1.000 – 1.081 |
| 8 | Karnataka | 1.103 | 1.002 – 1.275 |
| 9 | Punjab | 1.018 | 0.994 – 1.046 |
| 10 | Rajasthan | 0.995 | 0.931 – 1.058 |

### Phylogenomic analysis using maximum likelihood (MLE) tree construction

Maximum likelihood based phylogenetic tree construction using 564 genomes indicates clustering of entire dataset into two clades representing two distinct sub-lineages, BA.1 and BA.2 (Figure 2). In addition, this clustering was not found to be very tight and showed emergence of some sub-sub-lineages visible in the form of short and long branches radiating out within these clades.

**Figure 2.**
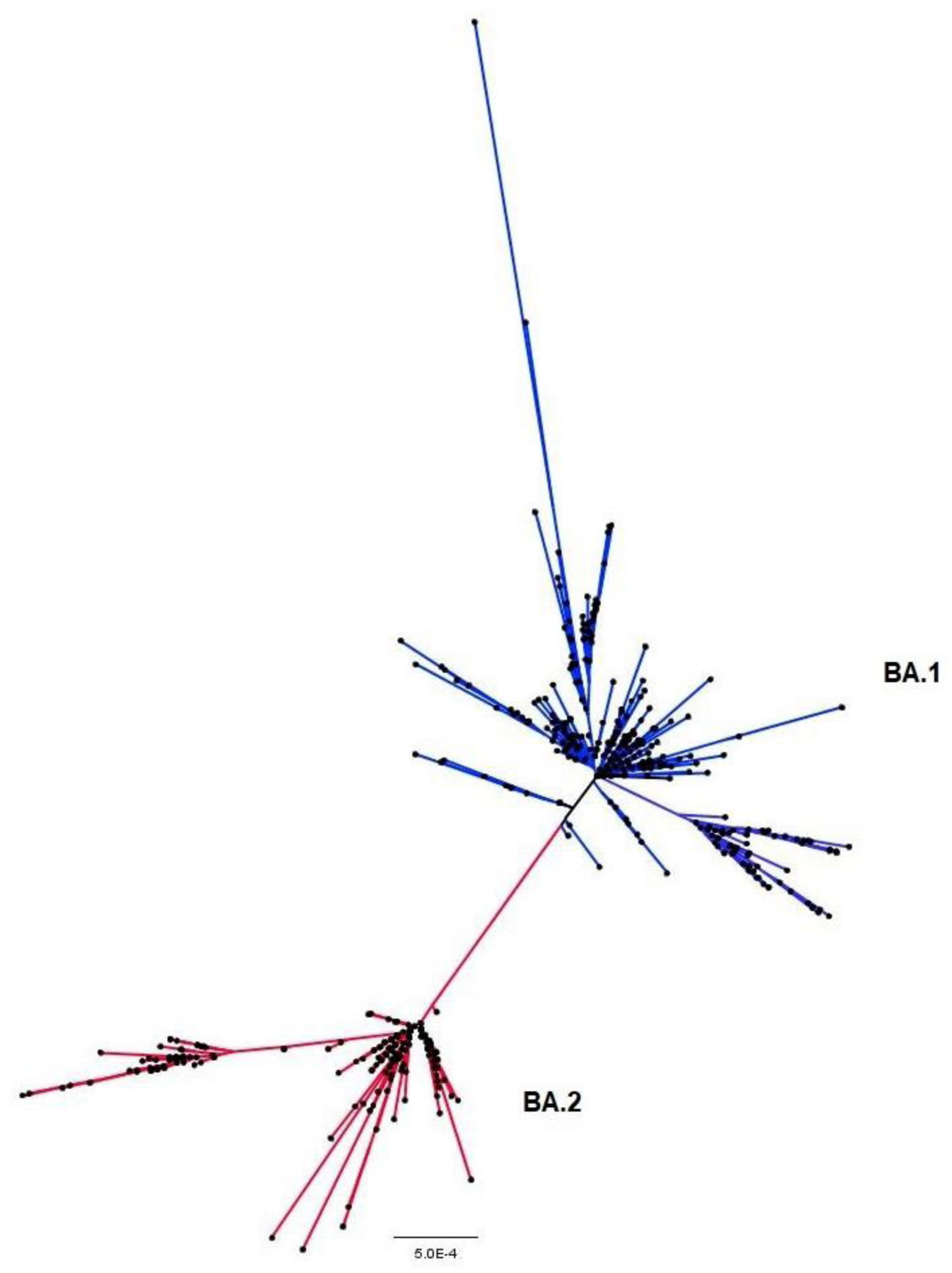
The Maximum likelihood tree constructed online PhyML showing clustering of 564 omicron genomes in two major clades. Note, the looseness of the cluster, which is characterized by short and long branches emerging as possible sub-sub lineages.

### Reduced Median (RM) Network construction

Based on the results of RM constructed from 297 haplotypes, eight major haplogroups, O1 to O8, were identified (Figure 3). Furthermore, these major haplogroups and their associated singleton haplotypes (shown by single color according to state) show interesting relatedness. Major and minor haplogroups as well as singleton haplotypes are connected through lineages defined by mutation numbers ranging between 1 to 9 (Figure 3). RM also reveals a total of over 30 median vectors in the network represented by black dots, which signify either unsampled, missing, or extinct taxa. The eight major haplogroups (O1-O8) are formed by the shared haplotypes between genomes of viruses distributed across a maximum of nine states to a minimum of five states. Moreover, there are equal numbers of minor haplogroups that results from the sharing of haplotypes between four and two states (not marked). A variant haplogroup marked as OA, represent unique ancestral position in the network because of its direct connection to the first omicron genome discovered in South Africa (Figure 3). Additionally, haplogroup O5 and O8 in the reconstructed RM are of particular interest since they acquired their haplotypes by sharing them across nine and eight states, respectively, and show broader distribution. Moreover, these two haplogroups (O5 and O8) exhibit a more star-like expansion, indicating the emergence of more autochthonous daughter haplogroups and haplotypes.

**Figure 3.**
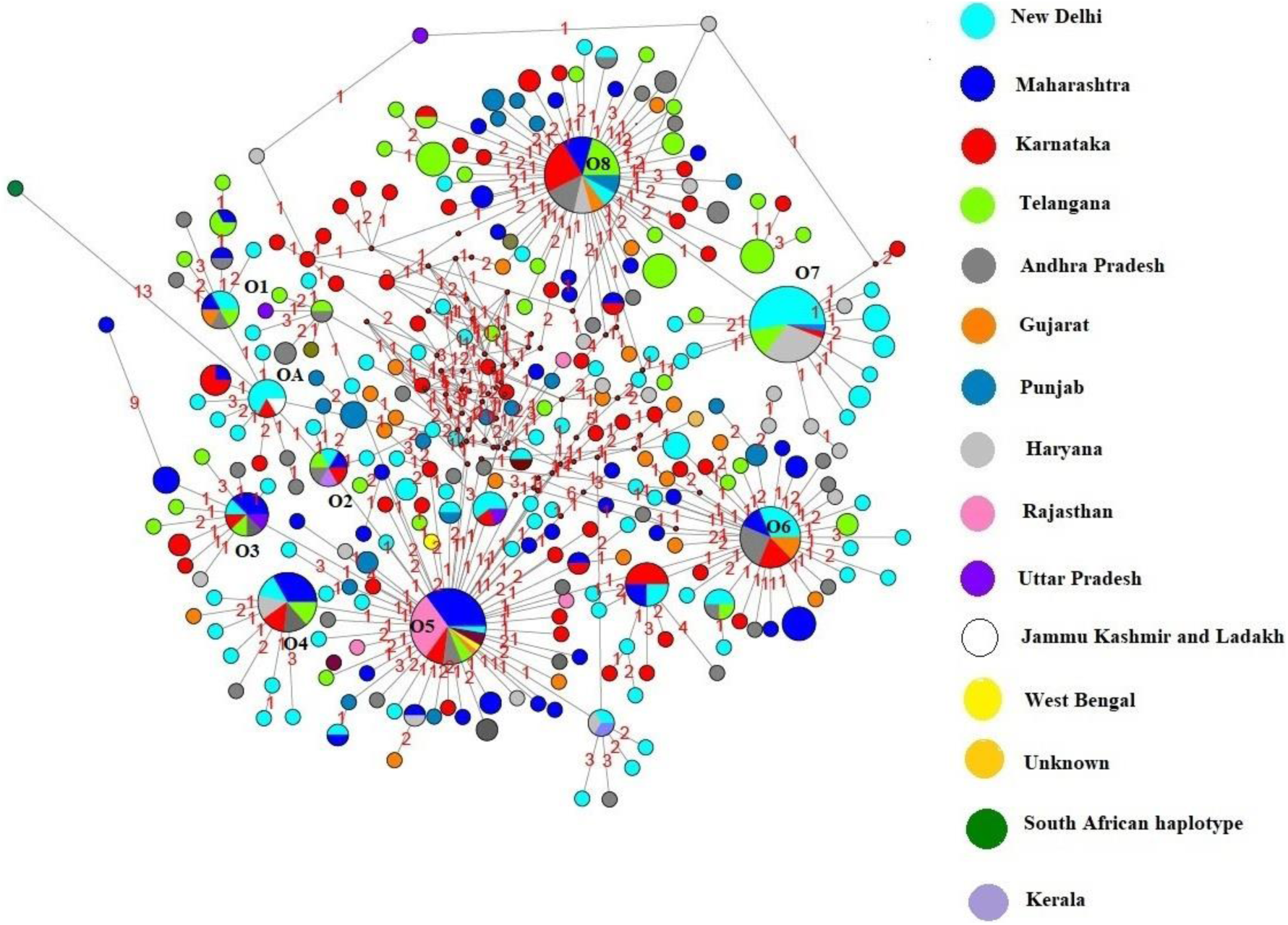
Reduced Median (RM) network constructed out of 297 omicron haplotypes representing various states of India. The major haplogroups inferred in the network are labelled O1-O8, while OA represents an ancestral haplogroup with connection to the early omicron cases detected in South Africa. The size of the circles corresponds to the number of shared haplotypes; Links with red numbers indicate number of mutated nucleotides. The black dots represent median vectors.

**Figure 4.**
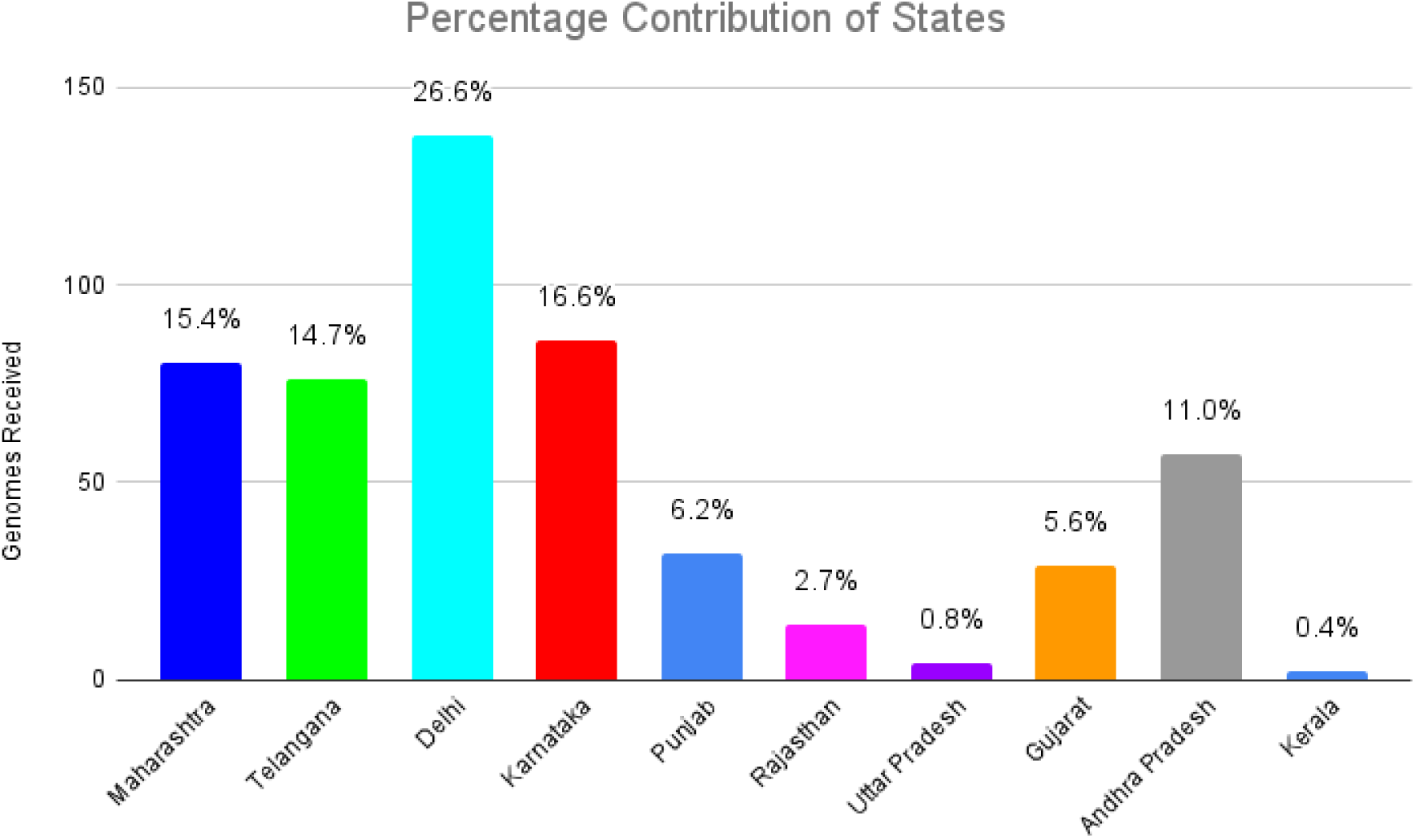
Percentage of genomes sequenced and deposited to GIASID from various states of India until 29^th^ January 2022.

## Discussion

The higher reproduction number (R0) of 2.445 for the BA.2 lineage is in line with [14] Lyngse et al. (2022) finding that BA.2 lineage is more transmissible in Denmark. Moreover, omicron variant exhibited higher recombination rates, facilitating rapid evolution [15]. The technical Advisory Group on SARS-CoV-2 Virus Evolution recommends public health authorities to monitor the BA.2 sub-lineage as a distinct sub-lineage of Omicron. Additionally, visibility of short and long branches within the BA.1 and BA2 clades inferred by maximum likelihood tree point towards recombination property of SARS-CoV-2 virus and their possibilities of evolving into new lineages (Fig 2). In addition to this, density of the population, vaccination status of the hosts, would also further determine the evolution of sub-sub lineages within the sub lineages into novel variants of concern. A study published by [16] Loconsole et al. (2022), reveals an outbreak of SARS-CoV-2 omicron infection and successful immune escape in 15 booster vaccinated healthcare workers in Italy. While study by [15] Liu et al. (2022) demonstrated chimerism in omicron variants, as a result of genomic recombination between two SARS-CoV-2 lineages involving the spike protein gene. Consequently, in this course of omicron evolution, emergence of new sub lineages and variants in future cannot be ruled out. Median Joining (MJ) and Reduced Median (RM) network enables geneticist to study genetic pattern in the context of relatedness among species or taxa. While used in viruses, organisms that are prone to recombination and horizontal gene transfer should be used with caution [17]. Rapid evolution of omicron variant as observed in the maximum likelihood tree is also supported by RM network constructed using same dataset. The higher number of major haplogroup including sub haplogroups and associated singleton haplogroups further confirms rapid evolutionary rate of recombination in both lineages (Fig 3). Haplogroups are generally regarded as relics of shared ancestry [18, 19]. Haplotype sharing has been used in contact tracing accurately at the beginning of the SARS-CoV-2 pandemic [20]. We propose that the information obtained from such networks can be explored in contact tracing, filling gaps, and community transmission in India. When we compared this RM with the first phylogenetic network published by [20] Forster et al. 2020 at the onset of the epidemic and with our own previously published MJ network [21], significant differences were observed. In comparison with our previously published MJ, RM exhibits a more distinct and faster evolution of variant in human hosts, evident in the emergence of more variants/haplogroups (colored circles) and the accumulation of large number of mutations across connecting lineages. We argue that the possible sub-sub lineages and several novel haplogroups inferred by our phylogenetic and RM network construction method respectively have the potential to be evolved into more transmissible and infectious variants. Thus, to monitor SARS-CoV-2 evolution in real time with the aim of developing more effective vaccines in the future, we recommend strict tracking of lineages through timely and regular genome sequencing of viruses across India.

## Conclusion

Our first-ever omicron genome- based study representing various states of India revel two lineages, BA.1 and BA.2. The BA.2 exhibits greater transmissibility. The study also shows the emergence of sub-sublineages and autochthonous haplogroups. In light of these findings, we recommend continuous and broad genome sequencing, and full vaccination of Indian population.

## Supporting information

Lineage Assignment

## Data Availability

The dataset used in this study is provided as supplementary material

## Acknowledgements

We gratefully acknowledge all authors and submitting laboratories of the sequences from GISAID.

