## Supplementary material for "Analyses of Omicron genomes from India reveal BA.2 as a more transmissible variant": Dataset

We gratefully acknowledge the following Authors from the Originating laboratories responsible for obtaining the specimens, as well as the Submitting laboratories where the genome data were generated and shared via GISAID, on which this research is based.

All Submitters of data may be contacted directly via www.gisaid.org

Authors are sorted alphabetically.

| Accession ID | Originating Laboratory | Submitting Laboratory | Authors |
|---|---|---|---|
| EPI_ISL_8285616 | ADUGODI DISPENSARY | National Center for Biological Sciences, TIFR - Rockefeller Foundation | Awadhesh Pandit; Bhagyashree Madhav Shelar; Darshan Sreenivas; Dimple Notani; Lakshminarayanan CP; Manisha Bharadwaj; Satyajit Mayor; Uma Ramakrishnan |
| EPI_ISL_8637800, EPI_ISL_8637814, EPI_ISL_8637817, EPI_ISL_9166224, EPI_ISL_9232037, EPI_ISL_9232038, EPI_ISL_9232039, EPI_ISL_9232044, EPI_ISL_9232045, EPI_ISL_9232046, EPI_ISL_9232048, EPI_ISL_9232060, EPI_ISL_9232071, EPI_ISL_9232086, EPI_ISL_9232088, EPI_ISL_9232089, EPI_ISL_9232090, EPI_ISL_9232091, EPI_ISL_9232097, EPI_ISL_9232102, EPI_ISL_9232108, EPI_ISL_9232130, EPI_ISL_9232136, EPI_ISL_9232146, EPI_ISL_9232154, EPI_ISL_9232155, EPI_ISL_9232156, EPI_ISL_9232160, EPI_ISL_9232161, EPI_ISL_9232163, EPI_ISL_9232168, EPI_ISL_9232178, EPI_ISL_9232179, EPI_ISL_9232256, EPI_ISL_9232266, EPI_ISL_9232278, EPI_ISL_9232279, EPI_ISL_9232308 | | | |
| see above | AP SSO | CSIR-Centre for Cellular and Molecular Biology-INSACOG | Amareshwar Vodapalli; Ara Sreenivas; Archana Bharadwaj Siva; B Himasri; Divya Tej Sowpati; Jandhyala Sai Krishna; Karthik Bharadwaj Tallapaka; Lamuk Zaveri; Malini Nemalikanti; Priya Nurkurthy; Rakesh K Mishra; Shreekant Verma; Sreelekshmi MS; Sumedha Avadhanula; Surabhi Srivastava; Tulasi Nagabandi; Valli Nagalakshmi Undamatla; Vidhyadhari Methuku |
| EPI_ISL_9175910, EPI_ISL_9175911, EPI_ISL_9175996, EPI_ISL_9175998, EPI_ISL_9175999, EPI_ISL_9176000, EPI_ISL_9176001, EPI_ISL_9176002 | | | |
| see above | BBMP- Mahadevapura | inStem, NCBS-INSACOG | Aswin SaiNarain; Dasaradhi Palakodeti; Uma Ramakrishnan |
| EPI_ISL_9175915, EPI_ISL_9175931, EPI_ISL_9175932, EPI_ISL_9175935, EPI_ISL_9175936, EPI_ISL_9175937, EPI_ISL_9175938, EPI_ISL_9175939, EPI_ISL_9175940, EPI_ISL_9176034, EPI_ISL_9176041, EPI_ISL_9176043, EPI_ISL_9176045, EPI_ISL_9176046 | | | |
| see above | BBMP- West | inStem, NCBS-INSACOG | Aswin SaiNarain; Dasaradhi Palakodeti; Uma Ramakrishnan |
| EPI_ISL_9175916, EPI_ISL_9175919, EPI_ISL_9175920, EPI_ISL_9175921, EPI_ISL_9175923, EPI_ISL_9175925, EPI_ISL_9175926, EPI_ISL_9175927 | | | |
| see above | BBMP- Yelahanka | inStem, NCBS-INSACOG | Aswin SaiNarain; Dasaradhi Palakodeti; Uma Ramakrishnan |
| EPI_ISL_9176003, EPI_ISL_9176009, EPI_ISL_9176011, EPI_ISL_9176016, EPI_ISL_9176018, EPI_ISL_9176019, EPI_ISL_9176020, EPI_ISL_9176022, EPI_ISL_9176023 | | | |
| see above | BBMP-Dasarahalli | inStem, NCBS-INSACOG | Aswin SaiNarain; Dasaradhi Palakodeti; Uma Ramakrishnan |
| EPI_ISL_9175909 | BBMP-Mahadevapura | inStem, NCBS-INSACOG | Aswin SaiNarain; Dasaradhi Palakodeti; Uma Ramakrishnan |
| EPI_ISL_9236277 | BJ Medical College, Ahmedabad | Gujarat Biotechnology Research Centre | Akhilesh Modi; Apurvasinh Puvar; Bhadreshsinh Gohil; Chaitanya Joshi; Disha Vora; Janvi Raval; Jaykumar Rangani; Madhvi Joshi; Nimesh Patel; Nitin Savaliya; NitinShukla; Priyank Chavda; Ramesh Pandit; Roshani Mishra; Sonal Sharma; Sumita Soni; Tasnim Trivedi; Zarna Patel |
| EPI_ISL_9175941, EPI_ISL_9175943, EPI_ISL_9175946, EPI_ISL_9175947, EPI_ISL_9175949, EPI_ISL_9175953, EPI_ISL_9175954, EPI_ISL_9175955, EPI_ISL_9175956, EPI_ISL_9175957, EPI_ISL_9175959, EPI_ISL_9175960, EPI_ISL_9175961, EPI_ISL_9175964, EPI_ISL_9175965, EPI_ISL_9175967, EPI_ISL_9175970, EPI_ISL_9175971, EPI_ISL_9175973, EPI_ISL_9175978, EPI_ISL_9175979, EPI_ISL_9175981, EPI_ISL_9175982, EPI_ISL_9175983, EPI_ISL_9175984, EPI_ISL_9175985, EPI_ISL_9175988, EPI_ISL_9175989, EPI_ISL_9175991, EPI_ISL_9175992, EPI_ISL_9175993 | | | |
| see above | Bangalore Airport | inStem, NCBS-INSACOG | Aswin SaiNarain; Dasaradhi Palakodeti; Uma Ramakrishnan |
| EPI_ISL_9176024, EPI_ISL_9176025, EPI_ISL_9176026, EPI_ISL_9176028, EPI_ISL_9176030, EPI_ISL_9176031, EPI_ISL_9176032 | | | |
| see above | Bengaluru Rural | inStem, NCBS-INSACOG | Aswin SaiNarain; Dasaradhi Palakodeti; Uma Ramakrishnan |
| EPI_ISL_8109017 | CHC Dakor | Gujarat Biotechnology Research Centre | Akhilesh Modi; Apurvasinh Puvar; Bhadreshsinh Gohil; Chaitanya Joshi; Disha Vora; Janvi Raval; Jaykumar Rangani; Madhvi Joshi; Nimesh Patel; Nitin Savaliya; Nitin Shukla; Priyank Chavda; Ramesh Pandit; Roshani Mishra; Sonal Sharma; Tasnim Trivedi; Zarna Patel |
| EPI_ISL_8769739 | CNC PATH LAB | ILBS | Arjun Bhugra; Chhagan Bihari Sharma; Ekta Gupta; Pramod Gautam; Rahul Garg; Reshu Agarwal; Shiv Kumar Sarin; Urvinder Kaur; Varun Suroliya |
| EPI_ISL_8769645 | CORE Diagnostics Private Limited | ILBS | Arjun Bhugra; Chhagan Bihari Sharma; Ekta Gupta; Pramod Gautam; Rahul Garg; Reshu Agarwal; Shiv Kumar Sarin; Urvinder Kaur; Varun Suroliya |
| EPI_ISL_8637640 | CSIR-National Environmental Engineering Research Institute | CSIR-Centre for Cellular and Molecular Biology-INSACOG | Amareshwar Vodapalli; Ara Sreenivas; Archana Bharadwaj Siva; B Himasri; Divya Tej Sowpati; Karthik Bharadwaj Tallapaka; Krishna Khairnar; Lamuk Zaveri; Malini Nemalikanti; Priya Nurkurthy; Rakesh K Mishra; Shreekant Verma; Sreelekshmi MS; Sumedha Avadhanula; Surabhi Srivastava; Tulasi Nagabandi; Valli Nagalakshmi Undamatla; Vidhyadhari Methuku |
| EPI_ISL_9023203 | Cellcare Patholab, Ahmedabad | Gujarat Biotechnology Research Centre | Akhilesh Modi; Apurvasinh Puvar; Bhadreshsinh Gohil; Chaitanya Joshi; Dhyanesh Desai; Disha Vora; Janvi Raval; Jaykumar Rangani; Madhvi Joshi; Nimesh Patel; Nitin Savaliya; Nitin Shukla; Priyank Chavda; Ramesh Pandit; Roshani Mishra; Sonal Sharma; Tasnim Trivedi; Zarna Patel |
| EPI_ISL_9232398 | Continental Hospital | CSIR-Centre for Cellular and Molecular Biology | Amareshwar Vodapalli; Ara Sreenivas; Archana Bharadwaj Siva; B Himasri; Divya Tej Sowpati; Jandhyala Sai Krishna; Karthik Bharadwaj Tallapaka; Lamuk Zaveri; Malini Nemalikanti; Priya Nurkurthy; Rakesh K Mishra; Shreekant Verma; Sreelekshmi MS; Sumedha Avadhanula; Surabhi Srivastava; Tulasi Nagabandi; Valli Nagalakshmi Undamatla; Vidhyadhari Methuku |
| EPI_ISL_7623676 | Dept of Microbilogy, Maulana Azad Medical College | J6094_MAMC_LNH | Avinash Lomash; Mohammad Faruq; Oves Siddiqui; SCOG_MAMC_LNH; Seema Kapoor. Sunil Kumar Polipalli; Somesh Kumar; Suresh Kumar |
| EPI_ISL_8868207 | Dept of Microbiology, Maulana Azad Medical College | GENOME SEQUENCING LABORATARY, Dept of Pediatrics, LOK NAYAK Hospital | Arvind Mohan; Avinash Lomash; INSACOG; Meenakshi Bothra; Mohammed Faruq; Oves Siddiqui; Prashanth N Suravajhala; Seema Kapoor.Sandeep Garg; Somesh Kumar; Sonal Saxena; Sunil Kumar Polipalli; Suresh Kumar; Vikas Manchanda |
| EPI_ISL_7644473 | Dept of Microbiology, Maulana Azad Medical College & Genome Sequencing Lab, Division of Genetics, Department of Pediatrics, Maulana Azad Medical College & Associated Lok Nayak Hospital | Genome Sequencing Lab, Division of Genetics, Department of Pediatrics, Maulana Azad Medical College & Associated Lok Nayak Hospital | Avinash Lomash; Mohammad Faruq; Oves Siddiqui; SCOG_MAMC_LNH; Seema Kapoor. Sunil Kumar Polipalli; Somesh Kumar; Suresh Kumar |
| EPI_ISL_9234081, EPI_ISL_9234082 | GMERS Medical College, Valsad | Gujarat Biotechnology Research Centre | Akhilesh Modi; Apurvasinh Puvar; Bhadreshsinh Gohil; Chaitanya Joshi; Disha Vora; Janvi Raval; Jaykumar Rangani; Madhvi Joshi; Nimesh Patel; Nitin Savaliya; NitinShukla; Priyank Chavda; Ramesh Pandit; Roshani Mishra; Sonal Sharma; Tasnim Trivedi; Vicky Gandhi; Zarna Patel |
| EPI_ISL_8380408, EPI_ISL_8380410, EPI_ISL_8452449, EPI_ISL_8452453, EPI_ISL_8452455, EPI_ISL_8452456, EPI_ISL_8452458, EPI_ISL_8452459, EPI_ISL_8452464, EPI_ISL_8452465, EPI_ISL_8452466, EPI_ISL_8452467, EPI_ISL_8452468, EPI_ISL_8452469, EPI_ISL_8452470, EPI_ISL_8452471, EPI_ISL_8452472, EPI_ISL_8452473, EPI_ISL_8452478, EPI_ISL_8452480, EPI_ISL_8452492, EPI_ISL_8452493, EPI_ISL_8452494, EPI_ISL_8452496, EPI_ISL_8452497, EPI_ISL_8452498, EPI_ISL_8452500, EPI_ISL_8452501, EPI_ISL_8452502, EPI_ISL_8452506, EPI_ISL_8452507, EPI_ISL_8452508, EPI_ISL_8527502, EPI_ISL_8527509, EPI_ISL_8527512, EPI_ISL_8527524, EPI_ISL_8527529, EPI_ISL_8527533, EPI_ISL_8527536, EPI_ISL_8527537, EPI_ISL_8527542, EPI_ISL_8527543, EPI_ISL_8527548, EPI_ISL_8527550, EPI_ISL_8527552, EPI_ISL_8527554, EPI_ISL_8527564, EPI_ISL_8527568, EPI_ISL_8527571, EPI_ISL_8527572, EPI_ISL_8527573, EPI_ISL_8527574, EPI_ISL_8527575, EPI_ISL_8527578, EPI_ISL_8527579, EPI_ISL_8527580, EPI_ISL_8527585, EPI_ISL_8527586, EPI_ISL_8527589, EPI_ISL_8527591 | | | |
| see above | Gandhi Medical College and Hospital (GMCH), Secunderabad | NIV Influenza | D.R.Manisha Rani; Dr.G.Sushma Rajya Lakshmi; Dr.K.Nagamani; Dr.Sunitha Pakalapaty |
| EPI_ISL_8830460, EPI_ISL_8830479 | GenePath, Pune | RF-IISER Pune | Aurnab Ghose; Joy Merwin Monteiro; Krishanpal Karmodiya; Nikhil Phadke; Shatakshi Ranade |
| EPI_ISL_9236268 | General Hospital, Lunavada, Mahisagar | Gujarat Biotechnology Research Centre | Akhilesh Modi; Apurvasinh Puvar; Bhadreshsinh Gohil; C.K.Solanki; Chaitanya Joshi; Disha Vora; Janvi Raval; Jaykumar Rangani; Madhvi Joshi; Nimesh Patel; Nitin Savaliya; NitinShukla; Priyank Chavda; Ramesh Pandit; Roshani Mishra; Sonal Sharma; Tasnim Trivedi; Zarna Patel |
| EPI_ISL_9234083 | Green Cross Genetics Lab Pvt Ltd, Ahmedabad | Gujarat Biotechnology Research Centre | Akhilesh Modi; Apurvasinh Puvar; Bhadreshsinh Gohil; Chaitanya Joshi; Disha Vora; Janvi Raval; Jaykumar Rangani; Madhvi Joshi; Nimesh Patel; Nitin Savaliya; NitinShukla; Priyank Chavda; Ramesh Pandit; Roshani Mishra; Sonal Sharma; Tasnim Trivedi; Umang Patel; Zarna Patel |
| EPI_ISL_8885401 | Gujarat Pathology laboratory & Diagnostic Center, Paldi, Ahmedabad | Gujarat Biotechnology Research Centre | Akhilesh Modi; Apurvasinh Puvar; Bhadreshsinh Gohil; Chaitanya Joshi; Disha Vora; Janvi Raval; Jaykumar Rangani; Madhvi Joshi; Nimesh Patel; Niraj Kotha; Nitin Savaliya; NitinShukla; Priyank Chavda; Ramesh Pandit; Roshani Mishra; Sonal Sharma; Tasnim Trivedi; Zarna Patel |
| EPI_ISL_8769564 | House of Diagnostics | ILBS | Arjun Bhugra; Chhagan Bihari Sharma; Ekta Gupta; Pramod Gautam; Rahul Garg; Reshu Agarwal; Shiv Kumar Sarin; Urvinder Kaur; Varun Suroliya |
| EPI_ISL_8445176, EPI_ISL_8445224, EPI_ISL_8445225, EPI_ISL_8445227, EPI_ISL_8445229, EPI_ISL_8445230, EPI_ISL_8445234, EPI_ISL_8445236, EPI_ISL_8445237, EPI_ISL_8445238, EPI_ISL_8445239, EPI_ISL_8445240, EPI_ISL_8445241, EPI_ISL_8445242, EPI_ISL_8445248, EPI_ISL_8572639, EPI_ISL_8572640, EPI_ISL_8572642, EPI_ISL_8572644, EPI_ISL_8572645, EPI_ISL_8572647, EPI_ISL_8572648, EPI_ISL_8572649, EPI_ISL_8572650, EPI_ISL_8572651, EPI_ISL_8572652, EPI_ISL_8572654, EPI_ISL_8572655, EPI_ISL_8572656, EPI_ISL_8572657, EPI_ISL_8572658, EPI_ISL_8572660, EPI_ISL_8572662 | | | |
| see above | ICMR-National Institute of Virology - INSACOG | NIV Influenza | Dr. Varsha Potdar and NIC Team |
| EPI_ISL_7381102, EPI_ISL_8542829, EPI_ISL_8542830, EPI_ISL_8542831, EPI_ISL_8542832, EPI_ISL_8542833, EPI_ISL_8542834, EPI_ISL_8542836, EPI_ISL_8542837, EPI_ISL_8542838, EPI_ISL_8542840, EPI_ISL_8542841, EPI_ISL_8542844, EPI_ISL_8542845, EPI_ISL_8542848, EPI_ISL_8542849, EPI_ISL_8542850, EPI_ISL_8542851, EPI_ISL_8542852, EPI_ISL_8542853, EPI_ISL_8542855, EPI_ISL_8542858, EPI_ISL_8542859, EPI_ISL_8542860, EPI_ISL_8542861, EPI_ISL_8542863, EPI_ISL_8542864, EPI_ISL_8542865, EPI_ISL_8542869, EPI_ISL_8542871, EPI_ISL_8542872, EPI_ISL_8542873, EPI_ISL_8542875, EPI_ISL_8542879, EPI_ISL_8542881, EPI_ISL_8542882, EPI_ISL_8542883, EPI_ISL_8542884, EPI_ISL_8542885, EPI_ISL_8542888, EPI_ISL_8542889, EPI_ISL_8542892, EPI_ISL_8542894, EPI_ISL_8542895, EPI_ISL_8542897, EPI_ISL_8542898, EPI_ISL_8542899, EPI_ISL_8542900, EPI_ISL_8542901, EPI_ISL_8542905, EPI_ISL_8542906, EPI_ISL_8542913, EPI_ISL_8542916, EPI_ISL_8542919, EPI_ISL_8542926, EPI_ISL_8542930, EPI_ISL_8542931, EPI_ISL_8542932, EPI_ISL_8542934, EPI_ISL_8542935, EPI_ISL_8542936, EPI_ISL_8542937, EPI_ISL_8542938, EPI_ISL_8542939, EPI_ISL_8542940, EPI_ISL_8542941, EPI_ISL_8542942, EPI_ISL_8542943, EPI_ISL_8542960, EPI_ISL_8603362 | | | |
| see above | Indian Council of Medical Research-National Institute of Virology, Microbial Containment Complex | Indian Council of Medical Research-National Institute of Virology, Microbial Containment Complex | Pragya D. Yadav |
| EPI_ISL_7986436, EPI_ISL_7986439, EPI_ISL_8712911, EPI_ISL_8712912, EPI_ISL_8712915, EPI_ISL_8712919, EPI_ISL_8712926, EPI_ISL_8712927, EPI_ISL_8712934, EPI_ISL_8712935, EPI_ISL_8712939, EPI_ISL_8712940, EPI_ISL_8712943, EPI_ISL_8712944, EPI_ISL_8712945, EPI_ISL_8712946, EPI_ISL_8712947, EPI_ISL_8712948, EPI_ISL_8712950, EPI_ISL_8712954, EPI_ISL_8712957, EPI_ISL_8712958, EPI_ISL_8712959, EPI_ISL_8712960, EPI_ISL_8712961, EPI_ISL_8712962, EPI_ISL_8712963, EPI_ISL_8712964, EPI_ISL_8712966, EPI_ISL_8712967, EPI_ISL_8712968, EPI_ISL_8712970, EPI_ISL_8712972, EPI_ISL_8712973, EPI_ISL_8712974, EPI_ISL_8712975, EPI_ISL_8712981, EPI_ISL_8712984, EPI_ISL_8712992, EPI_ISL_8712994, EPI_ISL_8712995, EPI_ISL_8712997, EPI_ISL_8713001, EPI_ISL_8713002, EPI_ISL_8713003, EPI_ISL_8713004, EPI_ISL_8713006, EPI_ISL_8713008, EPI_ISL_8713015, EPI_ISL_8713016, EPI_ISL_8713017, EPI_ISL_8713020, EPI_ISL_8713022, EPI_ISL_8713025, EPI_ISL_8713032 | | | |
| see above | Kasturba Hospital Molecular Lab - INSACOG | Kasturba hospital WGS | Jayanthi Shastri; Vidushi Chitalia |
| EPI_ISL_7864703 | Lok Kayak Hospital | Lok Kayak Hospital | Avinash Lomash; Mohammed FARUQ; Oves Siddiqui; Prashanth Suravajhala SURESH Kumar; Somesh Kumar; Sunil k Polipalli; seema Kapoor and SCOG_MAMC_LNH Lab |
| EPI_ISL_7877006, EPI_ISL_7877026, EPI_ISL_7877093, EPI_ISL_7877115, EPI_ISL_7877191, EPI_ISL_7877201, EPI_ISL_7877202, EPI_ISL_7877203, EPI_ISL_7877297, EPI_ISL_7889640, EPI_ISL_7889641 | | | |
| see above | Lok Nayak Hospital | Lok Nayak Hospital | ; Avinash Lomash; Lok Nayak Hospital; Mohammed FARUQ; Oves Siddiqui; Prashanth Suravajhala; SURESH Kumar; Somesh Kumar; Sunil k Polipalli; seema Kapoor and Genome Sequencing Lab |
| EPI_ISL_7876997 | Lok Nayak Hospital, Delhi | Lok Nayak Hospital, metro Station Central, Jawaharlal Nehru Marg, Near Delhi Gate, New Delhi, Delhi 110002 | Avinash Lomash; Lok Nayak Hospital; Mohammed FARUQ; Oves Siddiqui; Prashanth Suravajhala; SURESH Kumar; Somesh Kumar; Sunil k Polipalli; seema Kapoor and Genome Sequencing Lab |
| EPI_ISL_8285588 | MEDALL CLUMAX DIAGNOSTICS/BBMP | National Center for Biological Sciences, TIFR - Rockefeller Foundation | Awadhesh Pandit; Bhagyashree Madhav Shelar; Darshan Sreenivas; Dimple Notani; Lakshminarayanan CP; Manisha Bharadwaj; Satyajit Mayor; Uma Ramakrishnan |
| EPI_ISL_9021847, EPI_ISL_9021848, EPI_ISL_9021849, EPI_ISL_9021853, EPI_ISL_9021855, EPI_ISL_9021856, EPI_ISL_9021857, EPI_ISL_9021871, EPI_ISL_9021874, EPI_ISL_9021875, EPI_ISL_9021876, EPI_ISL_9021877, EPI_ISL_9021879, EPI_ISL_9021880, EPI_ISL_9021883, EPI_ISL_9021888, EPI_ISL_9021889, EPI_ISL_9021890, EPI_ISL_9021891, EPI_ISL_9021892, EPI_ISL_9021894, EPI_ISL_9021895 | | | |
| see above | Maharshi Dayanand University-Rohtak, CSIR-Institute of Genomics and Integrative Biology (CSIR-IGIB) | MDU, Rohtak, Haryana/CSIR-IGIB | Nar Singh Chauhan; Rajesh Pandey; Ranjeet Maurya; Varsha Ravi |
| EPI_ISL_9236266 | Manglam In Vitro Laboratory, Rajkot | Gujarat Biotechnology Research Centre | Akhilesh Modi; Apurvasinh Puvar; Bhadreshsinh Gohil; Chaitanya Joshi; Disha Vora; Janvi Raval; Jaykumar Rangani; Madhvi Joshi; Nimesh Patel; Nitin Savaliya; NitinShukla; Priyank Chavda; Rahul Thumbar; |

| | | | |
|---|---|---|---|
| | | | Ramesh Pandit; Roshani Mishra; Sonal Sharma; Tasnim Trivedi; Zarna Patel |
| EPI_ISL_8637669, EPI_ISL_8637811, EPI_ISL_8637838, EPI_ISL_9166255, EPI_ISL_9232081, EPI_ISL_9232100, EPI_ISL_9232101, EPI_ISL_9232105, EPI_ISL_9232106, EPI_ISL_9232126, EPI_ISL_9232128, EPI_ISL_9232141, EPI_ISL_9232184, EPI_ISL_9232185, EPI_ISL_9232186, EPI_ISL_9232231, EPI_ISL_9232232, EPI_ISL_9232285 | | | |
| see above | MapMyGenome | CSIR-Centre for Cellular and Molecular Biology-INSACOG | Amareshwar Vodapalli; Anuradha Acharya; Ara Sreenivas; Archana Bharadwaj Siva; B Himasri; Divya Tej Sowpati; Jandhyala Sai Krishna; Karthik Bharadwaj Tallapaka; Lamuk Zaveri; M. Aravind Kumar; Malini Nemalikanti; Priya Nurkurthy; Rakesh K Mishra; S.P.S. Satish; Sandhya Kiran; Shreekant Verma; Sreelekshmi MS; Sumedha Avadhanula; Surabhi Srivastava; Tulasi Nagabandi; Valli Nagalakshmi Undamatla; Vidhyadhari Methuku |
| EPI_ISL_8769742 | Medibird Diagnostics | ILBS | Arjun Bhugra; Chhagan Bihari Sharma; Ekta Gupta; Pramod Gautam; Rahul Garg; Reshu Agarwal; Shiv Kumar Sarin; Urvinder Kaur; Varun Suroliya |
| EPI_ISL_8494324, EPI_ISL_9234084, EPI_ISL_9234085 | Molecular lab, Shree Krishna Hospital, Karamsad, Anand | Gujarat Biotechnology Research Centre | Akhilesh Modi; Apurvasinh Puvar; Bhadreshsinh Gohil; Chaitanya Joshi; Chirag; Disha Vora; Janvi Raval; Jaykumar Rangani; Madhvi Joshi; Mustafa; Nimesh Patel; Nitin Savaliya; Nitin Shukla; NitinShukla; Priyank Chavda; Ramesh Pandit; Roshani Mishra; Sonal Sharma; Tasnim Trivedi; Zarna Patel |
| EPI_ISL_8769667, EPI_ISL_8769670 | NRL, Dr Lal Path Lab Ltds | ILBS | Arjun Bhugra; Chhagan Bihari Sharma; Ekta Gupta; Pramod Gautam; Rahul Garg; Reshu Agarwal; Shiv Kumar Sarin; Urvinder Kaur; Varun Suroliya |
| EPI_ISL_8881360, EPI_ISL_8881402, EPI_ISL_8881511, EPI_ISL_8881533, EPI_ISL_8881575, EPI_ISL_8881580, EPI_ISL_8881590, EPI_ISL_8881591, EPI_ISL_8881593, EPI_ISL_8881604, EPI_ISL_8881611, EPI_ISL_8881614, EPI_ISL_8881619, EPI_ISL_8881620, EPI_ISL_8881621, EPI_ISL_8881634, EPI_ISL_8881635, EPI_ISL_8881638, EPI_ISL_8881641, EPI_ISL_8881644, EPI_ISL_8881649, EPI_ISL_8881650, EPI_ISL_8881654, EPI_ISL_8881661, EPI_ISL_8881662, EPI_ISL_8881667, EPI_ISL_8881668, EPI_ISL_8881670, EPI_ISL_8881671, EPI_ISL_8881672, EPI_ISL_8881678, EPI_ISL_8881680, EPI_ISL_8881682, EPI_ISL_8881684, EPI_ISL_8881691, EPI_ISL_8881692, EPI_ISL_8881693, EPI_ISL_8881694, EPI_ISL_8881695, EPI_ISL_8881701, EPI_ISL_8881703, EPI_ISL_8881707, EPI_ISL_8881708, EPI_ISL_8881709, EPI_ISL_8881710, EPI_ISL_8881716, EPI_ISL_8881718, EPI_ISL_8881721, EPI_ISL_8881722, EPI_ISL_8881723, EPI_ISL_8881727, EPI_ISL_8881730, EPI_ISL_8881731, EPI_ISL_8881736, EPI_ISL_8881737, EPI_ISL_8881738, EPI_ISL_8881739, EPI_ISL_8881745, EPI_ISL_8881746, EPI_ISL_8881748, EPI_ISL_8881751, EPI_ISL_8881757, EPI_ISL_8881760, EPI_ISL_8881762, EPI_ISL_8881763, EPI_ISL_8881765, EPI_ISL_8881768, EPI_ISL_8881770, EPI_ISL_8881775, EPI_ISL_8881776, EPI_ISL_8881777, EPI_ISL_8881779, EPI_ISL_8881780, EPI_ISL_8881784, EPI_ISL_8881787, EPI_ISL_8881791, EPI_ISL_8881793, EPI_ISL_8881804, EPI_ISL_8881808, EPI_ISL_8881812, EPI_ISL_8881813, EPI_ISL_8881817, EPI_ISL_8881818, EPI_ISL_8881819, EPI_ISL_8881820, EPI_ISL_8881821, EPI_ISL_8881822, EPI_ISL_8881824, EPI_ISL_8881829, EPI_ISL_8881830, EPI_ISL_8881833, EPI_ISL_8881835, EPI_ISL_8881837, EPI_ISL_8881838, EPI_ISL_8881839, EPI_ISL_8881840, EPI_ISL_8881841, EPI_ISL_8881842, EPI_ISL_8881844, EPI_ISL_8881847, EPI_ISL_9003879 | | | |
| see above | National Centre for Disease control (NCDC), CSIR-Institute of Genomics and Integrative Biology (CSIR-IGIB) | NCDC/CSIR-IGIB/INSACOG | Aanchal Yadav; Anil Kumar; Aparna Swaminathan; Kriti Khare; Nisha Rawat; Pallavi Mishra; Partha Chattopadhyay; Priti Devi; Priyanka Mehta; Rajesh Pandey; Ranjeet Maurya; Shaista Praveen; Sheeba Saifi; Uzma Shamim; Varsha Ravi |
| EPI_ISL_8764350 | National Centre for Disease control (NCDC), CSIR-Institute of Genomics and Integrative Biology (CSIR-IGIB), Translational Health Science and Technology Institute (THSTI) | NCDC/CSIR-IGIB/THSTI | Aanchal Yadav; Anbalagan Ananthraj; Aparna Swaminathan; Guruprasad Medigeshi; Janmejey Singh; Kriti Khare; Pallavi Mishra; Partha Chattopadhyay; Priti Devi; Priyanka Mehta; Rajesh Pandey; Ranjeet Maurya; Shaista Praveen; Sheeba Saifi; Uzma Shamim; Varsha Ravi |
| EPI_ISL_9236270 | Nidan Path Lab, Gandhinagar | Gujarat Biotechnology Research Centre | Akhilesh Modi; Apurvasinh Puvar; Bhadreshsinh Gohil; Chaitanya Joshi; Disha Vora; Himani Patel; Janvi Raval; Jaykumar Rangani; Madhvi Joshi; Nimesh Patel; Nitin Savaliya; NitinShukla; Priyank Chavda; Ramesh Pandit; Roshani Mishra; Sonal Sharma; Tasnim Trivedi; Zarna Patel |
| EPI_ISL_8119542 | Northstar Pathology Lab, Ahmedabad | Gujarat Biotechnology Research Centre | Akhilesh Modi; Apurvasinh Puvar; Bhadreshsinh Gohil; Chaitanya Joshi; Disha Vora; Janvi Raval; Jaykumar Rangani; Jigar Suthar; Madhvi Joshi; Nimesh Patel; Nitin Savaliya; Nitin Shukla; Priyank Chavda; Ramesh Pandit; Roshani Mishra; Sonal Sharma; Tasnim Trivedi; Zarna Patel |
| EPI_ISL_9023145, EPI_ISL_9236271 | Northstar Pathology Laboratory, Ahmedabad | Gujarat Biotechnology Research Centre | Akhilesh Modi; Apurvasinh Puvar; Bhadreshsinh Gohil; Chaitanya Joshi; Disha Vora; Janvi Raval; Jaykumar Rangani; Kanupriya; Madhvi Joshi; Nimesh Patel; Nitin Savaliya; NitinShukla; Priyank Chavda; Ramesh Pandit; Roshani Mishra; Sonal Sharma; Tasnim Trivedi; Zarna Patel |
| EPI_ISL_8142222 | P.D.U Medical College Rajkot | Gujarat Biotechnology Research Centre | Akhilesh Modi; Apurvasinh Puvar; Bhadreshsinh Gohil; Chaitanya Joshi; Disha Vora; Janvi Raval; Jaykumar Rangani; Madhvi Joshi; Madhvlika Mistry; Nimesh Patel; Nitin Savaliya; Nitin Shukla; Priyank Chavda; Ramesh Pandit; Roshani Mishra; Sonal Sharma; Tasnim Trivedi; Zarna Patel |
| EPI_ISL_9236267 | Pathocare Pathology Laboratory, Vadodara | Gujarat Biotechnology Research Centre | Akhilesh Modi; Apurvasinh Puvar; Bhadreshsinh Gohil; Chaitanya Joshi; Disha Vora; Janvi Raval; Jaykumar Rangani; Madhvi Joshi; Nimesh Patel; Nitin Savaliya; NitinShukla; Priyank Chavda; Ramesh Pandit; Roshani Mishra; Sonal Sharma; Tasnim Trivedi; Viral Patel; Zarna Patel |
| EPI_ISL_9232416, EPI_ISL_9232419, EPI_ISL_9232422, EPI_ISL_9232426, EPI_ISL_9232428, EPI_ISL_9232429, EPI_ISL_9232435, EPI_ISL_9232437, EPI_ISL_9232447, EPI_ISL_9232448, EPI_ISL_9232449, EPI_ISL_9232454, EPI_ISL_9232455, EPI_ISL_9232457, EPI_ISL_9232459, EPI_ISL_9232464 | | | |
| see above | Rainbow Hospital | CSIR-Centre for Cellular and Molecular Biology | Amareshwar Vodapalli; Ara Sreenivas; Archana Bharadwaj Siva; B Himasri; Divya Tej Sowpati; Jandhyala Sai Krishna; Kalyan Ram Uppaluri; Karthik Bharadwaj Tallapaka; Lamuk Zaveri; Malini Nemalikanti; Priya Nurkurthy; Rakesh K Mishra; Shreekant Verma; Smita Juvvadi; Sreelekshmi MS; Sricharan Devineni; Sumedha Avadhanula; Surabhi Srivastava; Tulasi Nagabandi; Valli Nagalakshmi Undamatla; Vidhyadhari Methuku |
| EPI_ISL_9236274 | SSG Hospital Vadodara | Gujarat Biotechnology Research Centre | Akhilesh Modi; Apurvasinh Puvar; Bhadreshsinh Gohil; Chaitanya Joshi; Disha Vora; Janvi Raval; Jaykumar Rangani; Jigna Karia; Madhvi Joshi; Nimesh Patel; Nitin Savaliya; NitinShukla; Priyank Chavda; Ramesh Pandit; Roshani Mishra; Sonal Sharma; Tasnim Trivedi; Zarna Patel |
| EPI_ISL_8744182 | Scientific Diagnostic Centre Pvt. Ltd., Ahmedabad | Gujarat Biotechnology Research Centre | Akhilesh Modi; Apurvasinh Puvar; Bhadreshsinh Gohil; Chaitanya Joshi; Disha Vora; Janvi Raval; Jaykumar Rangani; Madhvi Joshi; Moksha Narechania; Nimesh Patel; Nitin Savaliya; NitinShukla; Priyank Chavda; Ramesh Pandit; Roshani Mishra; Sonal Sharma; Tasnim Trivedi; Zarna Patel |
| EPI_ISL_9234086 | Supratech Micropath Diagnostics & Research laboratory Ahmedabad | Gujarat Biotechnology Research Centre | Akhilesh Modi; Amee Shukla; Apurvasinh Puvar; Bhadreshsinh Gohil; Chaitanya Joshi; Disha Vora; Janvi Raval; Jaykumar Rangani; Madhvi Joshi; Nimesh Patel; Nitin Savaliya; NitinShukla; Priyank Chavda; Ramesh Pandit; Roshani Mishra; Sonal Sharma; Tasnim Trivedi; Zarna Patel |
| EPI_ISL_8494337 | Supratech Micropath Laboratory Research Institute Pvt Ltd, Ahmedabad | Gujarat Biotechnology Research Centre | Akhilesh Modi; Apurvasinh Puvar; Bhadreshsinh Gohil; Chaitanya Joshi; Disha Vora; Janvi Raval; Jaykumar Rangani; Madhvi Joshi; Nimesh Patel; Nitin Savaliya; Nitin Shukla; Priyank Chavda; Ramesh Pandit; Roshani Mishra; Shiva; Sonal Sharma; Tasnim Trivedi; Zarna Patel |
| EPI_ISL_9236272, EPI_ISL_9236273, EPI_ISL_9236276 | Symmers Pathocare Laboratory -Ahmedabad | Gujarat Biotechnology Research Centre | Akhilesh Modi; Apurvasinh Puvar; Bhadreshsinh Gohil; Chaitanya Joshi; Disha Vora; Janvi Raval; Jaykumar Rangani; Madhvi Joshi; Nimesh Patel; Nitin Savaliya; NitinShukla; Priyank Chavda; Ramesh Pandit; Roshani Mishra; Sonal Sharma; Tasnim Trivedi; Umang Gandhi; Zarna Patel |
| EPI_ISL_9092102, EPI_ISL_9092237, EPI_ISL_9092384, EPI_ISL_9092427 | TATA 1MG LABS - TATA 1MG Technologies Private Limited, Bengaluru | National Center for Biological Sciences, TIFR - Rockefeller Foundation | Awadhesh Pandit; Bhagyashree Madhav Shelar; Darshan Sreenivas; Dimple Notani; Kathirvel Kandasamy; Lakshminarayanan CP; Satyajit Mayor; Uma Ramakrishnan |
| EPI_ISL_9023165 | Unipath Speciality Laboaratory, Ahmedabad | Gujarat Biotechnology Research Centre | Akhilesh Modi; Apurvasinh Puvar; Bhadreshsinh Gohil; Chaitanya Joshi; Disha Vora; Janvi Raval; Jaykumar Rangani; Jwalant Shah; Madhvi Joshi; Nimesh Patel; Nitin Savaliya; NitinShukla; Priyank Chavda; Ramesh Pandit; Roshani Mishra; Sonal Sharma; Tasnim Trivedi; Zarna Patel |
| EPI_ISL_8925674 | Unipath Speciality Laboratory Limited, Ahmedabad | Gujarat Biotechnology Research Centre | Akhilesh Modi; Apurvasinh Puvar; Bhadreshsinh Gohil; Chaitanya Joshi; Disha Vora; Janvi Raval; Jaykumar Rangani; Jwalant Shah; Madhvi Joshi; Nimesh Patel; Nitin Savaliya; NitinShukla; Priyank Chavda; Ramesh Pandit; Roshani Mishra; Sonal Sharma; Tasnim Trivedi; Zarna Patel |
| EPI_ISL_9234080 | Universal Pathology & Molocular Lab, New Ranip, Ahmedabad | Gujarat Biotechnology Research Centre | Akhilesh Modi; Apurvasinh Puvar; Bhadreshsinh Gohil; Bipin Patel; Chaitanya Joshi; Disha Vora; Janvi Raval; Jaykumar Rangani; Madhvi Joshi; Nimesh Patel; Nitin Savaliya; NitinShukla; Priyank Chavda; Ramesh Pandit; Roshani Mishra; Sonal Sharma; Tasnim Trivedi; Zarna Patel |
| EPI_ISL_8801348, EPI_ISL_8801349, EPI_ISL_8801350, EPI_ISL_8801351, EPI_ISL_8801352, EPI_ISL_8801353, EPI_ISL_8801354, EPI_ISL_8801355, EPI_ISL_8801356, EPI_ISL_8801357, EPI_ISL_8801358, EPI_ISL_8801359, EPI_ISL_8801360, EPI_ISL_8801361, EPI_ISL_8801365, EPI_ISL_8801366, EPI_ISL_8801367, EPI_ISL_8801368, EPI_ISL_8801369, EPI_ISL_8801370, EPI_ISL_8801371, EPI_ISL_8801372, EPI_ISL_8801373, EPI_ISL_8801374, EPI_ISL_8801375, EPI_ISL_8801377, EPI_ISL_8801378, EPI_ISL_8801380 | | | |
| see above | VRDL,GMC,PTA | NIV Influenza | Dr.Rupinder Bakshi |
| EPI_ISL_8884441 | Viral Research and Diagnostic Laboratories | NIV Influenza | Dr.Rupinder Bakshi |
| EPI_ISL_9236269 | Zydus Hospital & Healthcare Research Pvt. Ltd. | Gujarat Biotechnology Research Centre | Akhilesh Modi; Apurvasinh Puvar; Bhadreshsinh Gohil; Chaitanya Joshi; Disha Vora; Janvi Raval; Jaykumar Rangani; Jaymin Patel; Madhvi Joshi; Nimesh Patel; Nitin Savaliya; NitinShukla; Priyank Chavda; Ramesh Pandit; Roshani Mishra; Sonal Sharma; Tasnim Trivedi; Zarna Patel |
